# Increase in SARS-CoV-2 seroprevalence in healthy blood donors after the second wave of COVID-19 pandemic in South-Eastern Italy: evidence for asymptomatic young individuals as potential virus spreaders

**DOI:** 10.1101/2021.07.12.21260358

**Authors:** Francescopaolo Antonucci, Josè Ramòn Fiore, Lucia De Feo, Tommaso Granato, Mariantonietta Di Stefano, Giuseppina Faleo, Ahmed Mohamed Farhan Mohamed, Maurizio Margaglione, Michele Centra, Teresa Antonia Santantonio

## Abstract

**Background:** Italy has been the first among western countries to experience SARS-CoV-2 spread during which the southern regions were also heavily affected by the pandemic. To understand and monitor properly the evolution of COVID-19 pandemic, population based seroprevalence studies are a valid tool for the infection rates and effective prevalence of the SARS-CoV-2.

**Aim:** In this prospective study, we assessed the changes in SARS-CoV-2 seroprevalence rates among non-vaccinated blood donors in South-Eastern Italy over May 2020 to March 2021.

**Methods:** 8,183 healthy blood donors referring to the Transfusion Center at the University Hospital “Riuniti” of Foggia (Italy) for blood donation in the period May 2020-March 2021 were tested for anti-SARS-CoV-2 antibodies by Ortho Clinical Diagnostics VITROS^®^ 3600. None of the considered subjects had a diagnosed symptomatic COVID-19 infection.

**Results:** Overall, 516 resulted positive for anti-SARS-CoV-2 IgG antibodies (6.3%, 95% CI, 0.03-0.15%), 387 (4.7%) were male and 129 (1.7%) female. A statistically significant increase in the seropositive population was found from May 2020 to March 2021 (Fisher’s p<0.001). The difference of the seroprevalence was significant in terms of age but not sex (2-sided p<0.05 for age; 2-sided p≥0.05 for sex) in both groups.

**Conclusion:** Our study shows a significant increase in the SARS-CoV-2 seroprevalence among blood donors and suggests a potential role of asymptomatic individuals in continuing the spread of the pandemic. These results may contribute to establishing containment measures and priorities in vaccine campaigns.

## INTRODUCTION

The Coronavirus Disease 2019 (COVID-19) pandemic resulted in more than 176 million cases and more than 3.5 million deaths worldwide as of June 13^th^, 2021 [1-2], putting a strain on the healthcare, economic and social system worldwide.

Italy has been the first among western countries to experience SARS-CoV-2 spread with a heterogeneous geographical distribution of the pandemic during the first wave (dramatic in the northern regions and more contained in the southern regions), but much more homogeneous in the second wave during which the southern regions were also heavily affected by the pandemic.

In June 2021, the number of COVID-19 infections resulted in more than 4 million cases in Italy [2], of which more than 250 thousand in Apulia (Southern Eastern Italy) [3]. A cumulative incidence of 625.6 per 10,000 inhabitants has been estimated in this region, and 6,545 deaths were recorded with a lethality rate of 2.6% [3].

The accurate assessment of antibodies seroprevalence during a pandemic can provide important information on pathogen exposure in the population [4-8]. In this regard, seroprevalence surveys in blood donors are recognized as valuable tools for refining estimates of infection and transmission [9-15].

In this prospective study, we assessed the anti-SARS-CoV-2 seroprevalence rates among non-vaccinated healthy blood donors in Foggia (South-Eastern Italy) over the period May 2020-March 2021.

## METHODS

### Study population

Between May 2020 and March 2021, 8,183 blood donors referring to the Transfusion Center at the “Ospedali Riuniti” University Hospital, Foggia (Italy) for blood donation were enrolled in the study. Exclusion criteria were active infections or medical conditions, reported risk factors for parenterally acquired infections, chronic degenerative conditions, diagnosis of cancer, or high risk of cardiovascular events. Clinical examination, medical history, and biochemical testing were performed on all donors. No recent symptoms COVID-19 related, nor close contact with confirmed cases or suspected cases had to be present in all subjects. None of the subjects had been vaccinated for SARS-CoV-2 infection.

Each blood donor signed an informed consent to be enrolled in the study.

### Detection of anti-SARS-CoV-2

Qualitative determination of serum total anti-Sars Cov 2 antibodies was performed as described elsewhere [16]. Briefly, the commercially available anti-SARS-CoV-2 Total reagent kit (Pencoed Bridgend, UK), a chemiluminescent immunoassay using mouse monoclonal human antibodies labelled with horseradish peroxidase (HRP) was used. Positive samples were then tested for the presence of specific IgG antibodies using the anti-SARS-CoV-2 IgG reagent kit (Pencoed Bridgend, UK).

### Statistical methods

Descriptive statistics was used to analyze the results. In particular, the Chi-square test and Fisher’s exact test were used to compare seropositive and seronegative donors’ characteristics and analyse the progressive increase of seroprevalence rates. Contingency tables were created to compare different donor characteristics. The estimated crude prevalence rate was adjusted according to the test’s sensitivity and specificity. All the analysis was performed using the SPSS software package (version 19.0) and Windows Excel^®^.

## RESULTS

Among the 8183 blood donors enrolled as a whole, 516 (6.3%) were tested positive for anti-SARS-CoV-2 IgG antibodies (Table 1). A statistically significant increase in the seropositive population was found from May 2020 to March 2021 (Figure 1) (Fisher’s p<0.001).

**Table 1:** Anti-Sars-CoV-2 seroprevalence in blood donors during the period May 2020-March 2021.

| Period<br>(Month/year) | Donors<br>(No) | Anti-Sars-CoV-2<br>seropositives<br>(No) | Crude prevalence rate for<br>seropositive donors<br>(%) |
| --- | --- | --- | --- |
| May 2020 | 883 | 9 | 1.0 |
| June '20 | 789 | 11 | 1.4 |
| July '20 | 645 | 10 | 1.6 |
| August '20 | 667 | 8 | 1.2 |
| September '20 | 901 | 12 | 1.3 |
| October '20 | 834 | 13 | 1.6 |
| November '20 | 560 | 38 | 6.8 |
| December '20 | 826 | 90 | 10.9 |
| January '21 | 796 | 85 | 10.7 |
| February '21 | 798 | 149 | 18.7 |
| March '21 | 484 | 91 | 18.8 |
| Total | 8183 | 516 | 6.3 |

**Figure 1:**
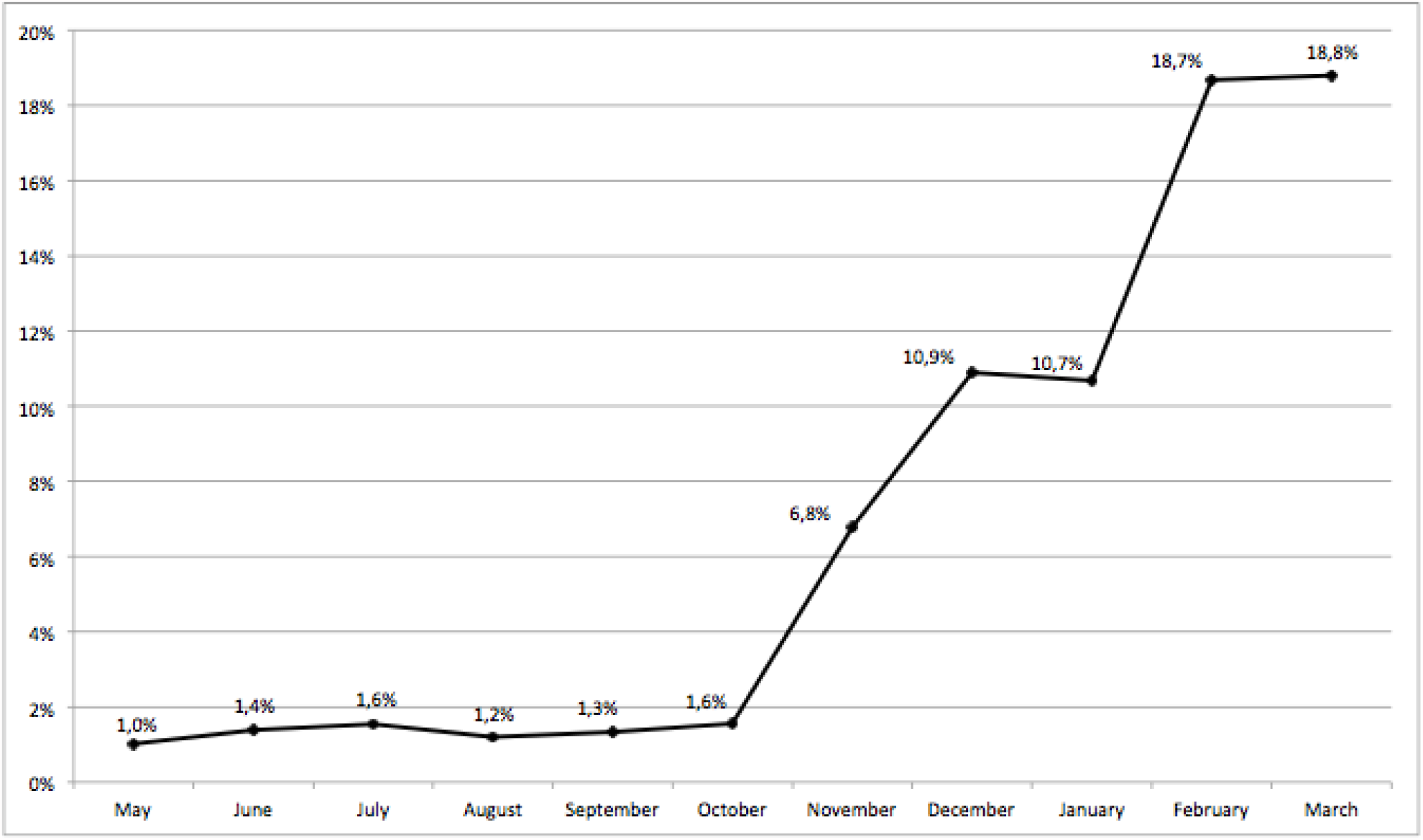
Cumulative percentage of seropositive blood donors in the period May 2020-March 2021.

Blood donors were divided into two groups by considering the donation period. Individuals in the first group were donors attending blood donation in May-September 2020, whereas blood donors of the second group were enrolled from October 2020 to March 2021. The first group included 4,018 people (2,958 men and 1,060 women, mean age 34 years), and the second group 4,165 people (3,178 men and 987 women, mean age 49 years) (Table 2). A statistically significant difference (2-sided p<0.001) of the seroprevalence rates was found between the two groups. The first group showed a seroprevalence of 0.5% (95% CI, 0.00-0.51%), while the second group showed a seroprevalence of 11.9% (95% CI, 0.08-0.10%). A higher percentage of seropositivity was detected in the second group in all age categories (Table 2).

**Table 2:** Demographic characteristics of 8183 blood donors tested for SARS-CoV-2 antibodies from May 2020 to September 2020 (Group 1) and from October 2020 to March 2021 (Group 2).

| Cts |  | Group 1 |  |  |  | Group 2 |  |  |  |
| --- | --- | --- | --- | --- | --- | --- | --- | --- | --- |
|  |  | Total | Seropos.<br>(No) | Seropos.<br>(%) | P-value | Total | Seropos.<br>(No) | Seropos.<br>(%) | P-value |
| Sex | M | 2958 | 12 | 0.4 | 0.30 | 3178 | 375 | 11.8 | 0.63 |
|  | F | 1060 | 7 | 0.6 |  | 987 | 122 | 12.4 |  |
| Age | 18-25 | 876 | 5 | 0.6 | 0.027 | 154 | 71 | 46.1 | <0.001 |
|  | 26-35 | 1410 | 13 | 0.9 |  | 218 | 97 | 44.5 |  |
|  | 36-45 | 1308 | 1 | 0.1 |  | 634 | 122 | 19.2 |  |
|  | 46-55 | 278 | 0 | 0 |  | 2113 | 131 | 6.2 |  |
|  | 56-65 | 138 | 0 | 0 |  | 981 | 75 | 7.6 |  |
|  | >65 | 8 | 0 | 0 |  | 65 | 1 | 1.5 |  |

Cts: Characteristics; F: Female; M: Male; Seropos.: Seropositives.

The difference of the seroprevalence was significant in terms of age but not sex (2-sided p<0.05 for age; 2-sided p>0.05 for sex) in both groups (Table 2).

None of the anti-SARS-CoV-2 positive subjects had a diagnosed symptomatic COVID-19 infection.

## DISCUSSION

In this study, we expanded a previous anti-SARS-CoV-2 seroprevalence survey among blood donors [17] and analysed 8,183 donors in the period May 2020-March 2021, including the first and the second waves of the COVID-19 pandemic in Italy.

In this large cohort of blood donors, we demonstrated a dramatic increase in terms of seroprevalence of anti-SARS-CoV-2 antibodies from 0.2% in May 2020 to 18.8% in March 2021.

Notably, none of the seropositive blood donors had been diagnosed as having experienced signs or symptoms of COVID-19 infection, and none had been subjected to vaccination. Thus, our survey provides a more accurate read of seroprevalence across subjects with asymptomatic infection, considered an essential source for SARS-CoV-2 transmission, but not detected by the national healthcare system.

The marked increase of seroprevalence observed over time is related to the progress of the pandemic in Italy.

After the first wave of the COVID-19 pandemic, Italy experienced very restricted containment measures for 69 days from March 9 to May 19, 2020, which dramatically reduced the COVID-19 burden in the whole country. The decreased number of new infections induced an unfounded optimism, which favoured a loss of adherence to preventive measures. Particularly during summer, relaxation of mobility reductions occurred, and several people traveled for vacation through regions with different prevalence rates, facilitating virus spread nationwide. Gathering on the beaches, crowded pubs, and discotheques were quite common during the “COVID-free” summer. All these behaviors had devastating effects in tourist areas and especially among young individuals.

Moreover, the reopening of schools in September probably contributed to a rapid increase in the number of infections among young individuals who are often asymptomatic but contagious, causing intra-family COVID-19 clusters.

Consequently, from September and with more evidence in October and November, a rapid spread in newly diagnosed infections was observed nationwide.

The analysis of the results in the two groups of blood donors, based on two different pandemic periods, clearly mirrors the evolution of the pandemic. The significant difference of the seroprevalence between Group 1 and 2 highlights the impact of containment strategies during the initial pandemic, which helped the reduction of SARS-CoV-2 circulation among the population, and confirm, in turn, the effects of the less restrictive containment actions applied in the summer period, that vanified such effects.

Apart from the general increase of blood donors positive for SARS-CoV-2 antibodies over time, we observed that the seroprevalence rate was low among all age categories in the first group. On the contrary, in the second group, the seroprevalence rate increased in all age categories, with the youngest groups showing the higher seropositive rates.

These data are of concern as young people have an active social life and may easily transmit the infection to others. To reduce the risk of transmission from asymptomatic people, measures such as wearing masks, hand hygiene, social distancing remain mandatory until individuals are widely immunized by vaccination. So far, young subjects have not been considered a priority population in vaccination campaigns. However, the critical role of young asymptomatic individuals in future SARS-CoV-2 spread, including viral variants, suggests a rapid extension of vaccination in this setting.

In conclusion, our study shows a significant increase in the SARS-CoV-2 seroprevalence among non-vaccinated blood donors and suggests a potential role of asymptomatic individuals in continuing the spread of the pandemic. The results of this study may contribute to establishing containment measures and priorities in vaccine campaigns.

## Data Availability

on request

